# A Sepsis Treatment Algorithm to Improve Early Antibiotic De-escalation While Maintaining Adequacy of Coverage (Early-IDEAS): A Prospective Observational Study

**DOI:** 10.1101/2022.04.13.22273851

**Authors:** Mohamed Bucheeri, Marion Elligsen, Philip W. Lam, Nick Daneman, Derek MacFadden

## Abstract

**Background:** Empiric antibiotic treatment selection should provide adequate coverage for potential pathogens while minimizing unnecessary broad-spectrum antibiotic use. We sought to pilot a rule- and model-based early sepsis treatment algorithm (Early-IDEAS) to make optimal individualized antibiotic recommendations.

**Methods:** The Early-IDEAS decision support algorithm was derived from previous Gram-negative and Gram-positive prediction rules and models. The Gram-negative prediction consists of multiple parametric regression models which predict the likelihood of susceptibility for each commonly used antibiotic for Gram-negative pathogens, based on epidemiologic predictors and prior culture results and recommends the narrowest spectrum agent that exceeds a predefined threshold of adequate coverage. The Gram-positive rules direct the addition or cessation of vancomycin based on prior culture results. We applied the algorithm to prospectively identified consecutive adults within 24-hours of suspected sepsis. The primary outcome was the proportion of patients for whom the algorithm recommended de-escalation of the primary antibiotic regimen. Secondary outcomes included: (1) the proportion of patients for whom escalation was recommended; (2) the number of recommended de-escalation steps along a pre-specified antibiotic cascade; and (3) the adequacy of therapy in the subset of patients with culture-confirmed infection.

**Results:** We screened 578 patients, of whom 107 eligible patients with sepsis were included. The Early-IDEAS treatment recommendation was informed by Gram-negative models in 76 (71%) of patients, Gram-positive rules in 66 (61.7%), and local guidelines in 27 (25%). Antibiotic de-escalation was recommended by the algorithm in almost half of all patients (n=50, 47%), no treatment change was recommended in 48 patients (45%), and escalation was recommended in 9 patients (8%). Amongst the patients where de-escalation was recommended, the median number of steps down the *a priori* antibiotic treatment cascade was 2. Among the 17 patients with relevant culture-positive blood stream infection, the clinician prescribed regimen provided adequate coverage in 14 (82%) and the algorithm recommendation would have provided adequate coverage in 13 (76%), p=1. Among the 25 patients with positive relevant (non-blood) cultures, the clinician prescribed regimen provided adequate coverage in 22 (88%) and the algorithm recommendation would have provided adequate coverage in 21 (84%), p=1.

**Conclusions:** An individualized decision support algorithm in early sepsis could lead to substantial antibiotic de-escalation without compromising adequate antibiotic coverage.

## INTRODUCTION

Antibiotic resistance is recognized as one of the greatest public health challenges of our time as it threatens the sustained availability of effective treatments for common infectious diseases^1^. Antibiotic use is thought to be the major driver of antibiotic resistance, and interventions to reduce unnecessary prescribing are urgently needed^2^.

However, as antibiotic resistance increases globally, it becomes more difficult to select and provide adequate empiric antibiotic therapy while complying with the principles of antibiotic stewardship, particularly in patients with life-threatening infections who stand to benefit the most from early adequate treatment^3,4^. Suspected sepsis is one of the most common indications in hospitals for the empiric use of antibiotics with a broad spectrum of activity given the high short-term mortality risk associated with this condition and the consequent fear of undertreatment that physicians often experience. However, the strategy of simply broadening the spectrum of empiric antibiotic treatment for all patients is not tenable as it favours the development of further resistance.

Current available evidence suggests that the use of computerized clinical decision support tools can increase the percentage of patients that receive desired care as was confirmed in a recent meta-analysis, but robust evidence on the use of this approach in patients with suspected sepsis is limited.^5^ Two previous studies indicated that clinical decision support algorithms including mathematical models and rules could be used to predict individual risk factors for resistance and successfully guide antibiotic selection.^6,7^ While effective, these approaches were limited by their focus on either Gram-positive or Gram-negative pathogens (separately), and were applied only after preliminary culture results were available. To improve on these, we have developed an encompassing algorithm, using modelling and rules, that considers all potential bacteria and can be used at the most critical empiric window (prior to microbiologic results). This early sepsis treatment algorithm could permit a narrower spectrum of therapy, while supporting the adequacy of coverage, allowing each patient to be on the right drug at the right time. This approach offers the potential for maximal benefits in adequate coverage and antibiotic stewardship by intervening earlier at the time of clinical presentation. However, early intervention poses additional challenges, and so our approach requires further validation before large-scale clinical implementation.

In this observational study, adult patients admitted at a large academic tertiary care center were prospectively reviewed to evaluate an early sepsis treatment algorithm. We hypothesized that the use of this new algorithm would have the potential to support early de-escalation recommendations while maintaining or improving time to adequate therapy.

## METHODS

### Study Setting, Design, and Participants

We performed a prospective observational study to evaluate the expected impact of an early sepsis treatment algorithm. This study was carried out at Sunnybrook Health Sciences Centre in Ontario, Canada, from November to December 2021. Ethics committee of Sunnybrook Research Institute has waived ethical approval for this work.

#### Patient Inclusion and Exclusion Criteria

The following inclusion criteria were used to identify adults with suspected early sepsis requiring empiric antibiotic treatment: (1) adult aged 18 years or older; (2) admitted to hospital; (3) received an eligible systemic antibiotic (See Figure 1); and (4) had blood cultures ordered within ±12 hours of receipt of antibiotics. The following exclusion criteria were applied: (1) patients already included in the study during prior episodes of early sepsis; (2) those receiving palliative care; (3) those who were pregnant; (4) other inpatient antibiotic use in the prior 72 hours; and (5) positive clinical culture associated within the index event available at the time of assessment. A roster of patients was generated twice daily during business hours Monday - Friday to identify patients that met the above criteria within the prior 24 hours. Patient charts were reviewed in real-time and the early sepsis treatment algorithm was applied to determine the recommended antibiotic regimen.

**Figure 1A.**
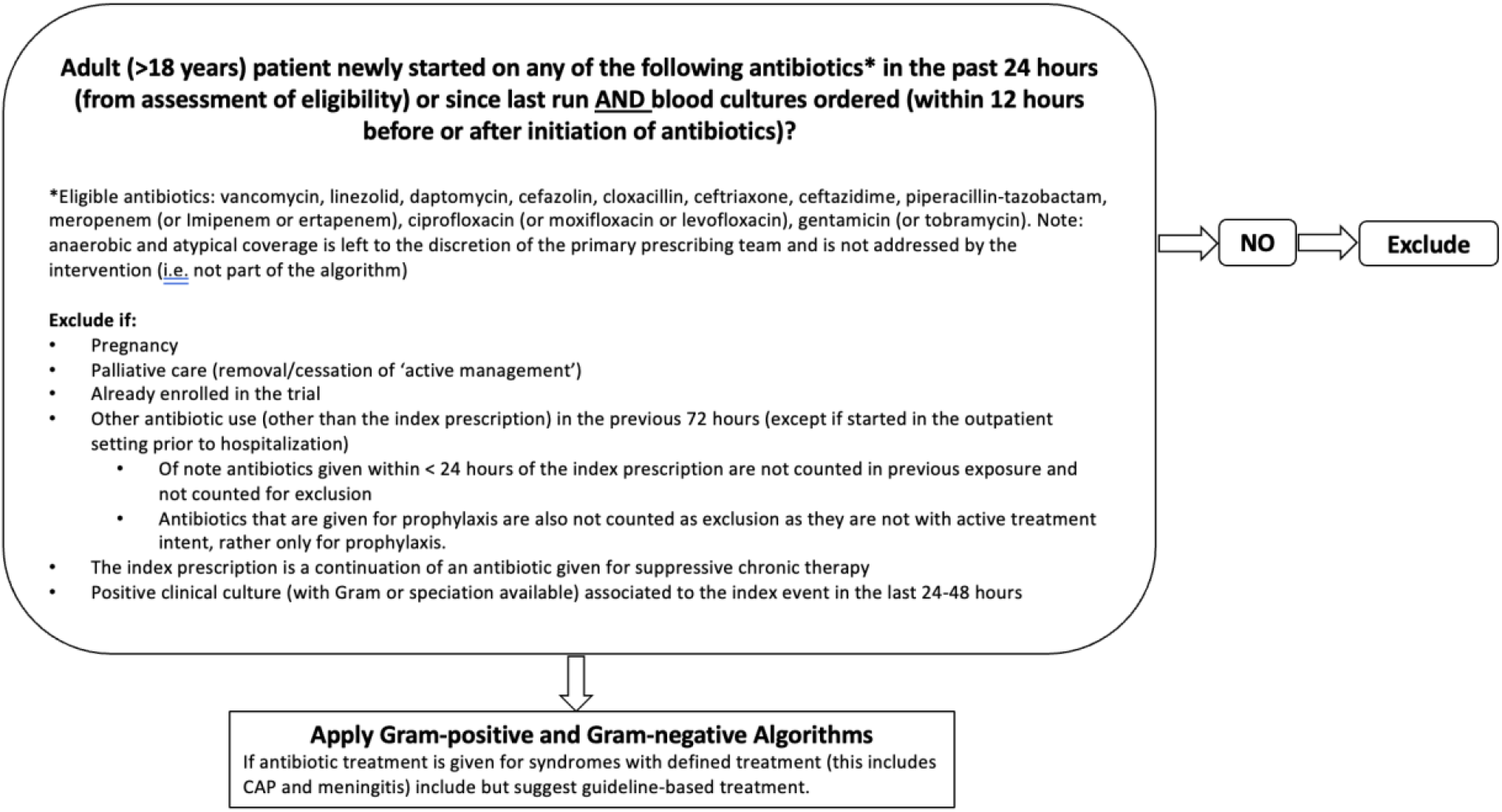
Eligibility for the algorithm.

**Figure 1B.**
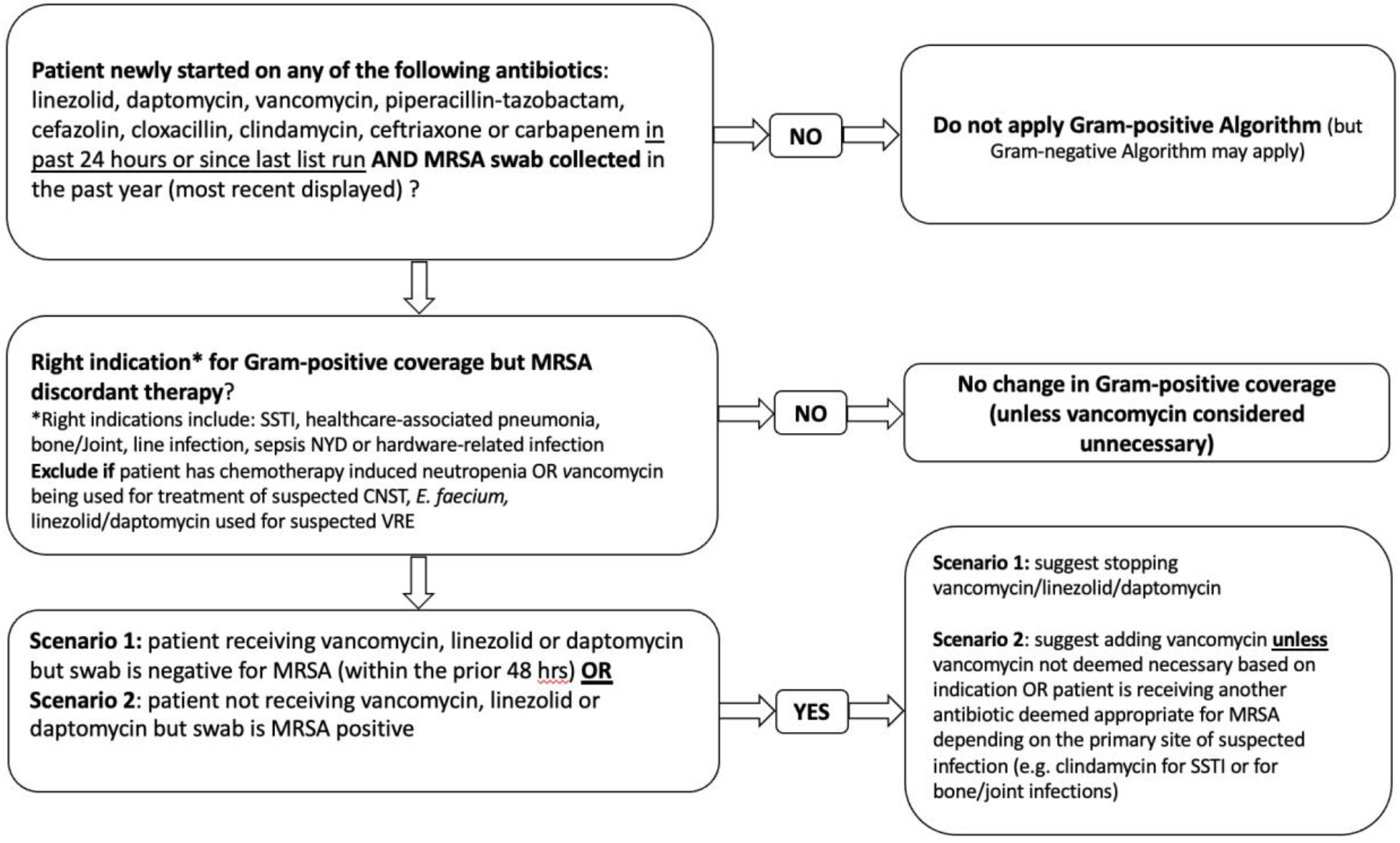
Gram-positive coverage component of the Early-IDEAS Algorithm.

**Figure 1C.**
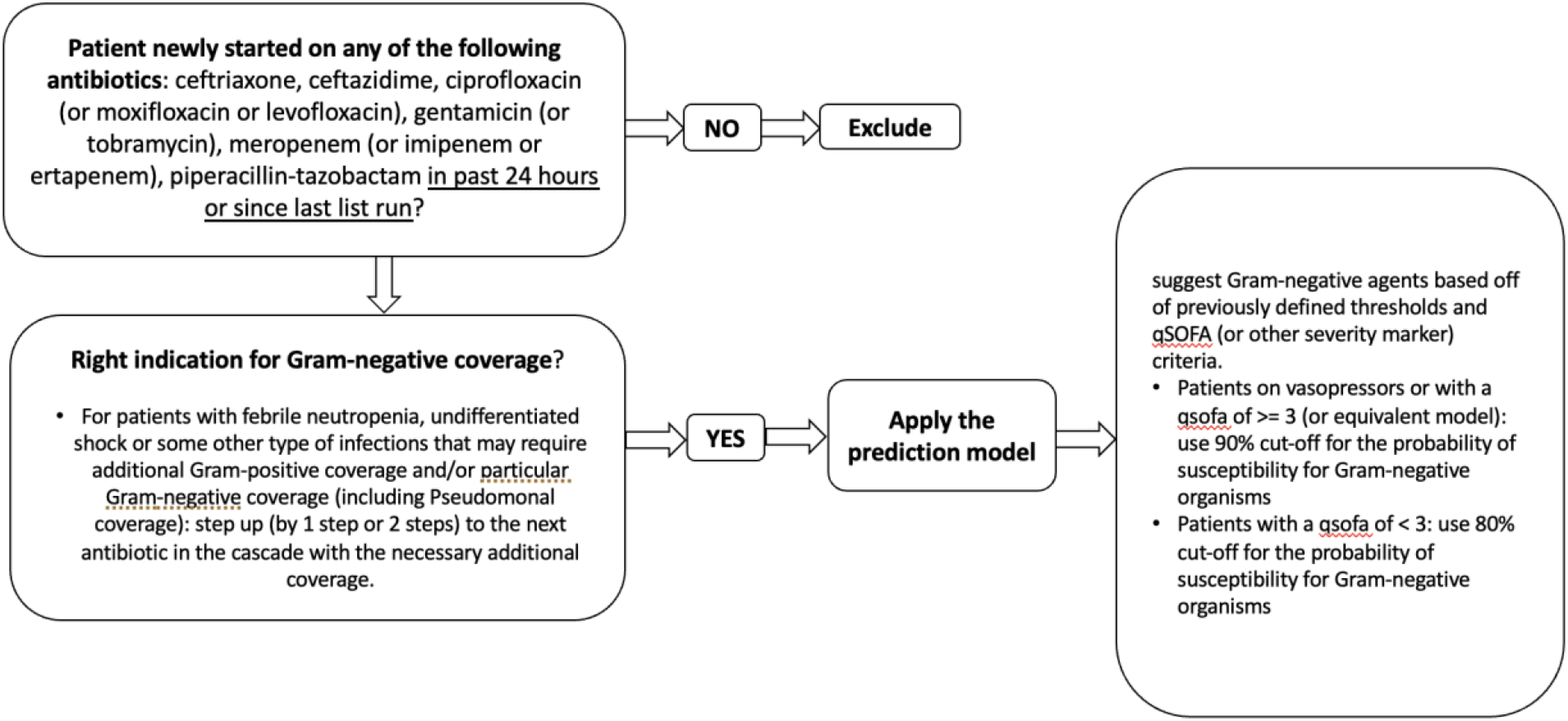
Gram-negative coverage component of the Early-IDEAS Algorithm.

### Early Sepsis Treatment Algorithm

We evaluated an early sepsis decision algorithm (Figure 1), which provided the empiric antibiotic selection in the context of suspected infection. This combined sepsis treatment algorithm was derived from rules and modelled prediction approaches that were developed and tested for both Gram-positive and Gram-negative infections^6,7^. The Gram-negative prediction model is as previously described^6^, and in brief consists of multiple parametric regression models which predict the likelihood of susceptibility for each commonly used antibiotic for Gram-negative pathogens, based on epidemiologic predictors (age, sex, prior hospitalization, prior ICU stay, prior antibiotic exposure) and prior culture results (prior antibiotic-resistant organisms from clinical cultures in the preceding year). These Gram-negative parametric models were validated and calibrated on historical culture data from the institution under study^6^. The Gram-positive algorithm is based on a previous prospective observational study and directs the addition or cessation of vancomycin based on prior culture results (Figure 1)^7^. For infections treated with a standard empiric regimen, regardless of patient risk factors, local guidelines for empiric therapy were applied (this included community-acquired pneumonia and meningitis). The treatment algorithm assumes an *a priori* antibiotic cascade for the treatment of Gram-negative pathogens, with the following antibiotics considered from broadest to narrowest respectively (meropenem>piperacillin-tazobactam>ceftazidime>ceftriaxone>ciprofloxacin). It seeks to move the prescriber down the antibiotic selection cascade to generally narrower spectrum agents by recommending the narrowest spectrum agent that still exceeds a pre-specified threshold of adequate coverage (80% for patients with a quick sequential organ failure assessment (qSOFA) score <2, 90% for patients with a qSOFA score > 2 or receiving vasopressor support))^8^. The purpose of this pilot study was to evaluate the potential impact of an early sepsis treatment algorithm, and therefore we did not provide treatment recommendations to the clinical team; the results of this pilot will inform further rigorous prospective evaluation.

### Outcomes and Predictor Variables

The main outcome measure of this study was the proportion of patients where de-escalation from the primary antibiotic regimen was recommended by our proposed approach. De-escalation was defined as movement down the aforementioned antibiotic cascade or the cessation of vancomycin or daptomycin. Escalation was defined as movement up the aforementioned antibiotic cascade or addition of vancomycin or daptomycin. Secondary outcomes that we evaluated included: (1) proportion of patients where escalation was recommended; (2) number of de-escalation steps along the antibiotic cascade that would be achieved; and (3) proportions of culture-positive patients who would receive adequate therapy with a given regimen (suggested vs. actual). Adequacy of therapy could only be determined for the subset of patients with a culture-positive infection. To describe the study population, and stratify by relevant factors, we collected the following predictor variables: age, sex, qSOFA score^9^, neutropenia, vasopressor use, mechanical ventilation, antibiotics prescribed at the time of assessment, clinical cultures, and prior Methicillin-resistant Staph. aureus (MRSA) screening cultures.

### Statistical Analysis

The primary and secondary outcomes were described using descriptive statistics. A comparison between the proportion of patients receiving adequate therapy for suggested versus received antibiotic therapy was performed using Fisher’s exact test. Some outcomes were stratified by relevant covariates.

## RESULTS

### Patient Characteristics

We reviewed 578 charts for eligibility during the study period, of which 471 were excluded (Figure 2). The majority of included patients received an antibiotic that had some degree of Gram-negative activity (92.5%), whereas only 8 patients received only Gram-positive active agents. The characteristics of the included patients are shown in Table 1. Less than half of the patients were female (n=42, 39.3%), the mean age was 67, and the majority (n=80, 74.8%) of patients had suspected community-acquired infection. Most patients were not markedly ill (median qSOFA score of 1), had undifferentiated sepsis (24.3%), and were most frequently prescribed piperacillin-tazobactam (n=47, 43.9%) for empiric therapy. These characteristics were relatively consistent across the patients who were de-escalated, escalated, or had no change to therapy.

**Table 1.**
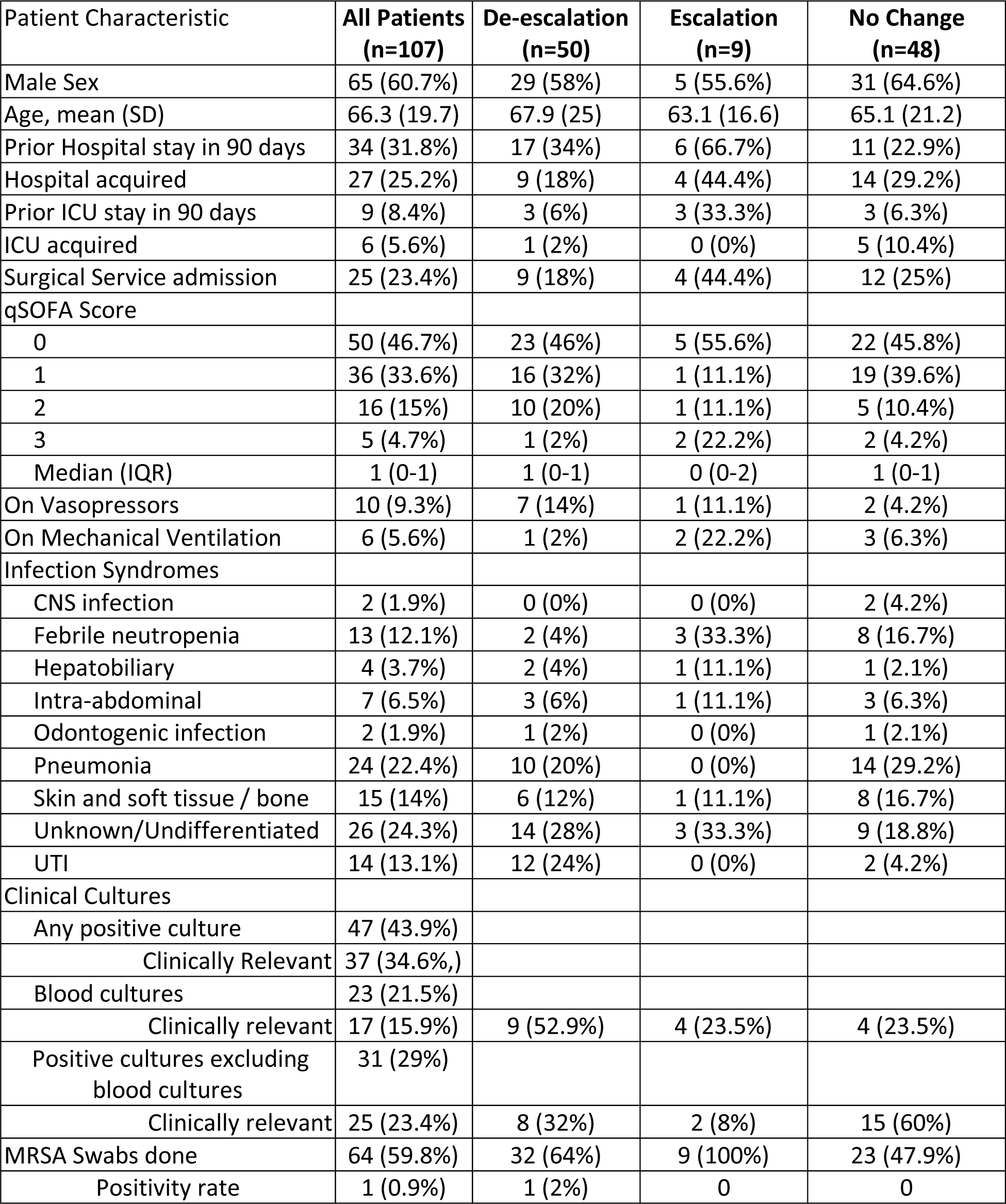
Characteristics of all patients (overall), and among those for whom the Early-IDEAS treatment algorithm recommended de-escalation, escalation, or no change in therapy

**Table 2.** Early-IDEAS algorithm components and antibiotic recommendations.

|  | All Patients | De-escalation<br>(n=50) | Escalation<br>(n=9) | No Change<br>(n=48) |
| --- | --- | --- | --- | --- |
| **Antibiotics Prescribed by Team** |  |  |  |  |
| Ampicillin | 1 (0.9%) | 0 (0%) | 0 (0%) | 1 (2.1%) |
| Azithromycin | 9 (8.4%) | 3 (6%) | 0 (0%) | 6 (12.5%) |
| Ciprofloxacin | 1 (0.9%) | 0 (0%) | 1 (11.1%) | 0 (0%) |
| Cefazolin | 10 (9.3%) | 0 (0%) | 1 (11.1%) | 9 (18.8%) |
| Ceftazidime | 1 (0.9%) | 1 (2%) | 0 (0%) | 0 (0%) |
| Ceftriaxone | 33 (30.8%) | 10 (20%) | 2 (22.2%) | 21 (43.8%) |
| Cephalexin | 1 (0.9%) | 0 (0%) | 1 (11.1%) | 0 (0%) |
| Clindamycin | 1 (0.9%) | 1 (2%) | 0 (0%) | 0 (0%) |
| Doxycycline | 2 (1.9%) | 0 (0%) | 0 (0%) | 2 (4.2%) |
| Ertapenem | 2 (1.9%) | 1 (2%) | 0 (0%) | 1 (2.1%) |
| Gentamicin | 1 (0.9%) | 0 (0%) | 0 (0%) | 1 (2.1%) |
| Levofloxacin | 1 (0.9%) | 0 (0%) | 0 (0%) | 1 (2.1%) |
| Metronidazole | 9 (8.4%) | 2 (4%) | 2 (22.2%) | 5 (10.4%) |
| Meropenem | 5 (4.7%) | 4 (8%) | 0 (0%) | 1 (2.1%) |
| Piperacillin-Tazobactam | 52 (48.6%) | 34 (68%) | 5 (55.6%) | 13 (27.1%) |
| Vancomycin | 11 (10.3%) | 5 (10%) | 0 (0%) | 6 (12.5%) |
| **Antibiotics Recommended by Algorithm** |  |  |  |  |
| Ampicillin | 1 (0.9%) | 0 (0%) | 0 (0%) | 1 (2.1%) |
| Azithromycin | 24 (22.4%) | 11 (22%) | 0 (0%) | 13 (27.1%) |
| Ciprofloxacin | 15 (14%) | 15 (30%) | 0 (0%) | 0 (0%) |
| Cefazolin | 9 (8.4%) | 0 (0%) | 0 (0%) | 9 (18.8%) |
| Ceftazidime | 1 (0.9%) | 0 (0%) | 0 (0%) | 1 (2.1%) |
| Ceftriaxone | 55 (51.4%) | 31 (62%) | 1 (11.1%) | 23 (47.9%) |
| Cephalexin | 0 (0%) | 0 (0%) | 0 (0%) | 0 (0%) |
| Clindamycin | 0 (0%) | 0 (0%) | 0 (0%) | 0 (0%) |
| Doxycycline | 0 (0%) | 0 (0%) | 0 (0%) | 0 (0%) |
| Ertapenem | 0 (0%) | 0 (0%) | 0 (0%) | 0 (0%) |
| Gentamicin | 0 (0%) | 0 (0%) | 0 (0%) | 0 (0%) |
| Levofloxacin | 0 (0%) | 0 (0%) | 0 (0%) | 0 (0%) |
| Metronidazole | 12 (11.2%) | 6 (12%) | 0 (0%) | 6 (12.5%) |
| Meropenem | 8 (7.5%) | 0 (0%) | 6 (66.7%) | 2 (4.2%) |
| Piperacillin-Tazobactam | 18 (16.8%) | 3 (6%) | 2 (22.2%) | 13 (27.1%) |
| Vancomycin | 8 (7.5%) | 2 (4%) | 0 (0%) | 6 (12.5%) |
| **Antibiotic step change** |  |  |  |  |
| 1 |  | 14 (28%) | 6 (66.7%) |  |
| 2 |  | 29 (58%) | 1 (11.1%) |  |
| 3 |  | 6 (12%) | 2 (22.2%) |  |
| 4 |  | 1 (2%) | 0 (0%) |  |
| Median (IQR) |  | 2 (1-2) | 1 (1-2) |  |
| **Guideline applied** | 27 (25.2%) | 8 (16%) | 0 (0%) | 19 (39.6%) |
| **Gram Negative Algorithm applied** | 76 (71%) | 42 (84%) | 9 (100%) | 25 (52.1%) |
| **Gram Positive model applied** | 66 (61.7%) | 30 (60%) | 9 (100%) | 27 (56.3%) |

**Figure 2.**
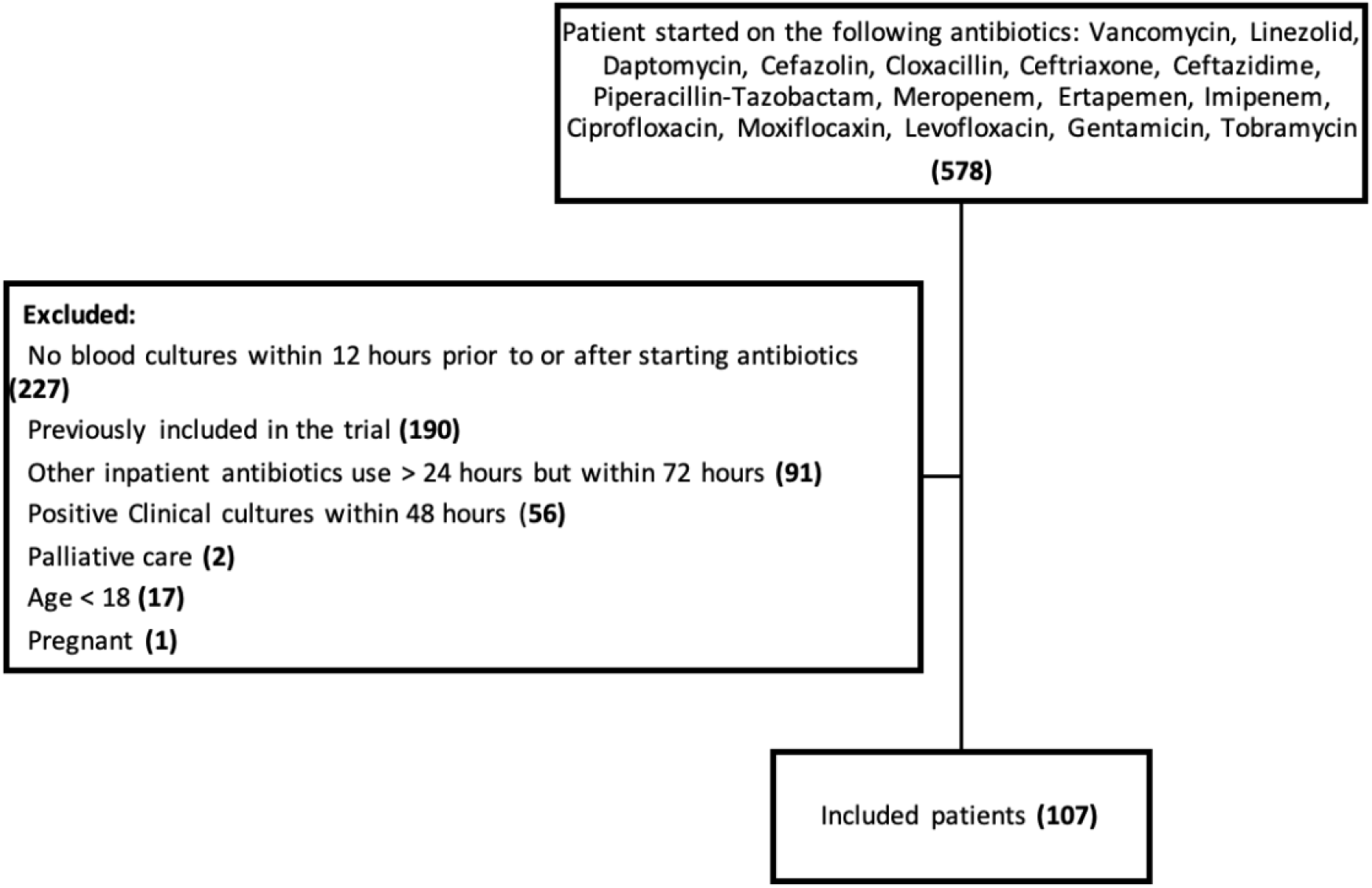
Study flow diagram.

Our treatment algorithm was applied to all eligible patients, and treatment recommendation was informed by the Gram-negative models in 76 patients (71%), by local guidelines in 27 (25.2%) and by Gram-positive rules in 66 (61.7%).

### Treatment Recommendations

Antibiotic de-escalation was recommended by the algorithm in almost half of all patients (n=50, 46.7%), no treatment change was recommended in 48 patients (44.9%), and escalation was recommended in 9 patients (8.4%). Amongst the patients where de-escalation was recommended, the median number of steps down the *a priori* antibiotic treatment cascade was 2. Many of these de-escalation steps involved the de-escalation of piperacillin-tazobactam to a narrower spectrum agent in the antibiotic cascade. In the small number of patients who were escalated, most of these (6/9, 66.7%) involved changing patients from piperacillin-tazobactam to meropenem due to a significant predicted risk for resistant Gram-negative pathogens.

### Adequacy of Coverage

A substantial subset of patients (n=47, 43.9%) was ultimately confirmed to have culture-positive infection whereby susceptibility results could be used to determine the adequacy of recommended coverage (Table 3). For patients with positive blood cultures, adequate antibiotics were prescribed by the clinical team in 14 (82.4%) patients. When applying the treatment algorithm for these same patients, adequate empiric antibiotic recommendations were similar, with 13 (76.5%) patients having adequate antibiotics recommended (p=1). For patients with positive (non-blood) clinical cultures, adequate antibiotics were prescribed in 22 (88%) patients. When applying the treatment algorithm for these same patients, adequate empiric antibiotic recommendations were again maintained, with 21 (84%) patients having adequate antibiotics recommended (p=1).

**Table 3.** Adequacy of antimicrobial coverage for a subset of patients with confirmed culture-positive infection.

|  | Patients with Adequate Empiric Antibiotic Coverage by Clinical Team | Patients with Adequate of Empiric Antibiotic Coverage by Early-IDEAS Algorithm |
| --- | --- | --- |
| **Blood Cultures Positive (n=17)\*** | 14 (82.4%) | 13 (76.5%) |
| Unchanged | 4 (28.6%) | 3 (23.1%) |
| De-escalated | 7 (50%) | 6 (46.2%) |
| Escalated | 3 (21.4%) | 4 (30.8%) |
| **Non-sterile site positive (n=25)\*\*** | 22 (88%) | 21 (84%) |
| Unchanged | 14 (63.6%) | 13 (61.9%) |
| De-escalated | 6 (27.3%) | 6 (28.6%) |
| Escalated | 2 (9.1%) | 2 (9.5%) |

\*Excludes 6 blood cultures with likely contaminants
\*\*Excludes 6 non-sterile site cultures (5 from un-affected body site, 1 likely contaminant)

## DISCUSSION

In this prospective validation of the study of the Early-IDEAS decision support algorithm we show that patients with suspected sepsis can be treated with narrower antibiotic agents while maintaining the adequacy of initial empiric coverage. The decision support algorithm we employed recommended de-escalation in almost half of all eligible patients, with a high proportion of blood and non-blood isolates adequately covered. Less than 1 in 10 patients had an escalation of antibiotics recommended. The Gram-negative prediction used, in brief, consists of multiple parametric regression models as discussed earlier. The treatment algorithm assumes an *a priori* antibiotic cascade for the treatment of Gram-negative pathogens and seeks to move the prescriber down the antibiotic selection cascade to generally narrower spectrum agents.

Clinical decision support tools for selecting antimicrobials in infectious syndromes are not new^10^; however, studies evaluating sepsis-specific support are less frequent. To date, there have been a number of studies evaluating support tools in the management of sepsis, though they are often focused on other (non-prescribing) aspects of management, including identifying prognosis and severity, determining likely discharge disposition, and the need for early treatment^11–13^. We did not identify any decision support tools developed for antibiotic prescribing in early sepsis that have been evaluated in a prospective randomized fashion, and there is a clear need for validated support models that can be tested in rigorous prospective trials.

Clinical decision support models, such as the Early-IDEAS treatment algorithm validated in this study, can be powered by institution-specific predictive models that have the ability to address the two competing aspects of antimicrobial stewardship: (1) the desire to retain high adequacy of empiric coverage while (2) reducing the breadth of antibiotic spectrum used in order to reduce the selection of resistance to reserve antimicrobial agents. Our approach fundamentally allows the provider to differentiate between patients that do not require broad-spectrum antibiotic therapy and those that do. Explicit, informed, and reproducible models have the potential to broadly support providers of all experience levels and backgrounds to operationalize antibiotic decision-making.

This study is intended as a pilot of early clinical decision support, and so is not powered to evaluate downstream clinical benefits of early de-escalation (such as reduction in antimicrobial costs, complications, and resistance) or the downstream benefits of adequate coverage (such as rapidity of clinical cure, reduced lengths of hospital stay and increased survival). The mathematical algorithms underpinning the decision support are derived from culture-positive infections, and we cannot be certain that culture-negative infections are caused by the same distribution of pathogens; this limitation is intrinsic to any antibiogram-based predictions.

Similarly, as anticipated only a subset of enrolled patients (34.6%) had microbiologically confirmed and clinically relevant pathogens, and so for the remainder, we are unable to compare the adequacy of coverage of the regimens recommended by the treating team versus our decision support algorithm. This is an expected trade-off of intervening early among patients presenting with sepsis. This pilot was designed to refine our algorithms and did not involve actual communication of treatment recommendations to the treating team, so we are not yet able to confirm whether clinicians will accept this decision support. However, prior work has suggested high uptake of recommendations^6,7^, and so we expect high uptake with future implementation of this decision support. Lastly, we found that concurrent MRSA swab results were generally not available at the early time points of assessment for patients during this pandemic period, and thus the Gram-positive arm of the algorithm was of limited utility for recommending cessation of vancomycin.

In summary, we demonstrate that a combined Gram-positive and Gram-negative decision support algorithm in early sepsis (Early-IDEAS) could help improve empiric antibiotic treatment by offering providers with narrower spectrum treatment options while maintaining high adequacy of therapy. This treatment algorithm requires further prospective evaluation to determine acceptability and efficacy. The time has come for the adoption of personalized medicine in sepsis treatment, and individualized models to support treatment selection will help us choose the right antibiotic at the right time.

## Data Availability

All data that can be made available for this present study is located within the manuscript.

